# A Stratified Worm Burden Framework for Endemic Equilibria, Demography, and Mass Drug Administration: Linking Classical Macroparasite Models to Transmission Dynamics

**DOI:** 10.64898/2026.10.08.26365093

**Authors:** D. Gurarie, CH King

**Affiliations:** Department of Mathematics, Applied Mathematics and Statistics, Center for Global Health and Diseases, CWRU, Cleveland, OH 44106; Center for Global Health and Diseases, School of Medicine, CWRU,, Cleveland, OH 44106

## Abstract

**Background:** Classical macroparasite models commonly represent worm burden heterogeneity through prescribed Poisson or negative-binomial (NB) distributions, while transmission dynamics are formulated in terms of aggregate quantities such as mean worm burden and prevalence. We revisit these assumptions using a stratified worm burden (SWB) framework that explicitly tracks the distribution of parasite burdens across heterogeneous host classes and allows demographic turnover, transmission, and interventions to be analyzed within a unified model.

**Methods:** We derive equilibrium distributions of the SWB system under homogeneous and heterogeneous infection processes and compare them with the classical Poisson and NB distributions used in macroparasite epidemiology and control policymaking. The framework is then extended to include host demography, coupled human-snail transmission dynamics, and periodic mass drug administration (MDA). Corresponding SWB and NB-type MacDonald models are calibrated to comparable endemic prevalence and mean worm burden, allowing direct comparison of potential transmission breakpoints and intervention outcomes.

**Results:** In the absence of demographic turnover, the SWB framework naturally generates Poisson and NB equilibrium distributions as limiting cases of homogeneous and heterogeneous infection processes. Incorporating demographic recruitment and mortality alters these benchmark distributions, producing endemic burden structures that differ systematically from their classical counterparts. Analysis of coupled human-snail systems reveals that transmission breakpoints and bistable equilibria occur only within a relatively restricted region of SWB parameter space. In contrast, comparable NB-MacDonald formulations exhibit breakpoint behavior over substantially broader parameter ranges with the net effect of overestimating the likelihood of interruption of transmission. Although calibrated models can reproduce near identical endemic prevalence and mean worm burden, they often generate markedly different responses to MDA, including differences in rebound dynamics, persistence, and elimination thresholds.

**Conclusions:** The SWB framework links classical statistical descriptions of worm aggregation to mechanistic burden-resolved population dynamics, clarifying how endemic distributions emerge and how they are modified by demography and control interventions. Our results suggest that transmission breakpoints are not a universal feature of helminth systems but depend strongly on model structure, and that predictions of elimination and post-treatment recovery may be considerably more model-dependent than implied by prevalence-based calibration alone. The SWB approach therefore provides a flexible alternative framework for evaluating parasite transmission dynamics and predicting the outcomes of control strategies.

## 1. Introduction

Mathematical models of macroparasite transmission have long provided the principal framework for understanding endemic infection dynamics and evaluating parasite control strategies. Classical formulations developed by Macdonald [1] and later extended by Anderson - May et al [2-6,10-11] describe infection in terms of mean worm burden (MWB), supplemented by assumed Poisson or negative-binomial (NB) worm-burden distributions to represent parasite aggregation among hosts. These models have generated many of the central concepts of helminth epidemiology, including endemic equilibria, mating effects, transmission thresholds, and elimination breakpoints.

A persistent challenge of MWB-based models is that worm-burden distributions are prescribed rather than derived from the underlying transmission process [9-11,16-17]. As a result, the connection between individual burden dynamics, population-level aggregation, demographic turnover, and intervention outcomes remains only partially resolved. These limitations become particularly important when evaluating long-term elimination strategies based on repeated mass drug administration (MDA), where treatment acts directly on individual worm burdens while model predictions are typically expressed through aggregate quantities such as prevalence and mean worm burden.

Stratified worm burden (SWB) models were developed to address these limitations by explicitly tracking the host population across worm-burden classes. Earlier work [6-7] demonstrated that SWB systems provide a natural framework for incorporating parasite aggregation and mating processes and can reproduce breakpoint phenomena traditionally associated with MWB models [1-5,16-17]. However, the breakpoint mechanism proposed in that analysis [7] relied on additional assumptions (Allee-effect) rather than arising directly from the underlying burden dynamics.

The present study revisits the basic SWB framework [7-8,20-23] that assumed a homogeneous host pool in terms of exposure (Poisson type SWB). Here we extend this framework to heterogeneous populations and show its connection to classical MacDonald models with overdispersal (Negative binomial-NB) burden distribution.

We then proceed to explore endemic equilibria and breakpoint behavior within a proper demographic makeup. We first show that Poisson and negative-binomial distributions emerge as equilibrium solutions of idealized SWB systems w/o population turnover, thereby providing a mechanistic foundation for the distributions commonly assumed in classical macroparasite theory. We then incorporate demographic recruitment and host turnover and demonstrate that realistic endemic burden distributions generally depart from these benchmark forms. Thus Poisson and NB distributions should be viewed as limiting reference states rather than generic endemic equilibria.

Building on these results, we compare MDA dynamics in the SWB framework with those of corresponding NB-type MacDonald systems calibrated to similar endemic prevalence and mean worm burden. Because treatment induces explicit redistribution among burden strata in SWB systems, agreement at endemic equilibrium does not necessarily imply agreement in post-treatment dynamics. We show that the two frameworks can differ substantially in rebound trajectories, persistence under repeated treatment, and predicted elimination outcomes.

We therefore revisit the SWB framework from three complementary perspectives. First, we establish Poisson and negative-binomial distributions as equilibrium solutions of idealized homogeneous and heterogeneous SWB systems, thereby providing a mechanistic interpretation of the statistical assumptions underlying classical macroparasite theory. Second, we examine how demographic recruitment and turnover modify these benchmark equilibria and generate endemic burden distributions that differ systematically from classical Poisson and NB forms. Third, we compare mass drug administration (MDA) dynamics and breakpoint behavior in SWB and corresponding negative-binomial MacDonald models calibrated to similar endemic prevalence and mean worm burden.

Our analysis shows that demographic structure, burden redistribution during treatment, and explicit worm-burden dynamics can substantially alter transmission equilibria and intervention outcomes. In particular, transmission breakpoints emerge only within a relatively restricted region of SWB parameter space, whereas comparable MacDonald formulations display bistability over much broader ranges. These findings suggest that elimination thresholds and post-treatment rebound may be more model-dependent than previously recognized.

## 2. Stratified Worm Burden (SWB) methodology

### 2.1 Standard version

The stratified worm burden (SWB) framework developed in [7-8,14,19-23] represents a host population as a set of worm-burden strata

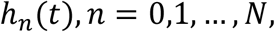

where *h*_*n*_ denotes the proportion of hosts carrying *n*adult worms at time *t*. The host population is normalized so that

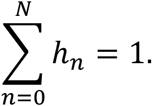

Hosts move between neighboring burden strata through worm acquisition and worm loss. The resulting population distribution provides a detailed description of infection intensity while retaining a relatively simple deterministic structure.

Let *λ* denote the mean rate of worm acquisition and *γ* the per-worm mortality rate.

For a homogeneous host population, worm acquisition increases burden by one worm,

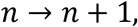

while worm mortality decreases burden by one worm,

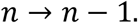

Figure 1 illustrates the burden lattice and transitions underlying the SWB formulation.

**Figure 1.**
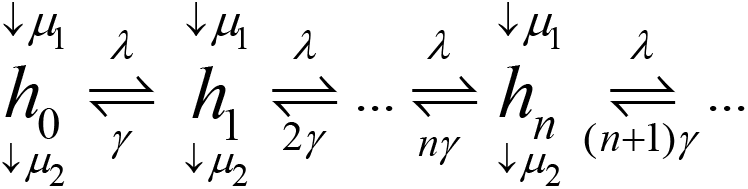
Structure of the stratified worm burden (SWB) model. Hosts are distributed among worm-burden strata h_n_. Worm acquisition moves hosts to higher burden classes, whereas worm mortality moves hosts to lower classes.

The resulting SWB dynamics can be written in matrix form

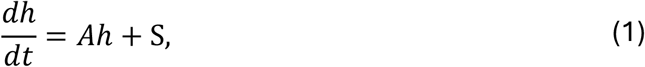

where *h* = (*h*_0_, *h*_1_, …, *h*_*N*_)^*T*^, *A* is the transition operator generated by acquisition and mortality processes, represented by tridiagonal matrix

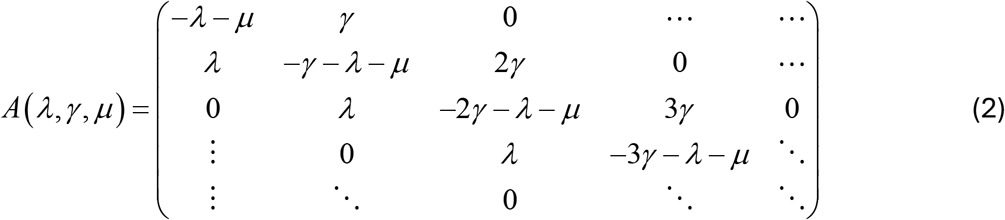

and *S* - demographic recruitment source. For child populations, recruitment enters primarily through the infection-free stratum *h*_0_, so *S* = (µ, 0, …, 0)^*T*^. Adult populations receive demographic inflow from maturing child cohorts and therefore inherit the infection structure of the younger population.

### 2.3 Mean worm burden and Equilibrium distribution

The mean worm burden is

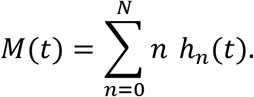

For the homogeneous SWB system, the governing equation for *M*can be obtained directly by taking the first statistical moment of the burden-stratified equations,

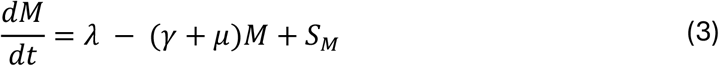

This relationship was derived previously and provides the connection between SWB and classical MacDonald mean-worm-burden (MWB) formulations.

Thus, SWB may be viewed as a distribution-based extension of MacDonald (MWB) system: whereas MWB tracks only the average infection level *M*, SWB follows the entire burden distribution *h* = (*h*_*n*_).

To derive the equilibrium distribution of SWB we consider the idealized case of infinitely many burden strata and no demographic source and turnover, µ = 0.The equilibrium state satisfies

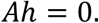

Its solution is the Poisson distribution with mean 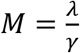

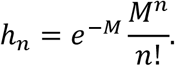

This result establishes the connection between homogeneous SWB and Poisson based classical MacDonald-type models (for details see Supplement).

## 3. Extended Risk-Heterogeneous SWB and Classical Equilibria

### 3.1 Distributed exposure risk

The standard SWB model assumes homogeneous host exposure and produces Poisson-type endemic equilibria. To account for the strong aggregation observed in helminth infections, we introduce a distributed exposure-risk factor.

Hosts are partitioned conceptually into risk classes *r* = 0, 1, 2, …, with geometrically distributed frequencies

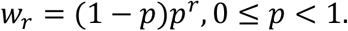

Base *p* characterizes the degree of exposure heterogeneity. Hosts in higher-risk classes acquire worms proportionally faster, *λ*_*r*_ = r *λ*, than the baseline acquisition rate *λ*.

Rather than introducing a second stratification level, we combine all risk classes carrying n worms into a single h_*n*_ stratum and modify the SWB transition operator accordingly.

The resulting model retains the original h_*n*_-stratification while introducing an additional parameter *p*. The schematic view of modified SWB in shown in Figure 2,

**Figure 2.**
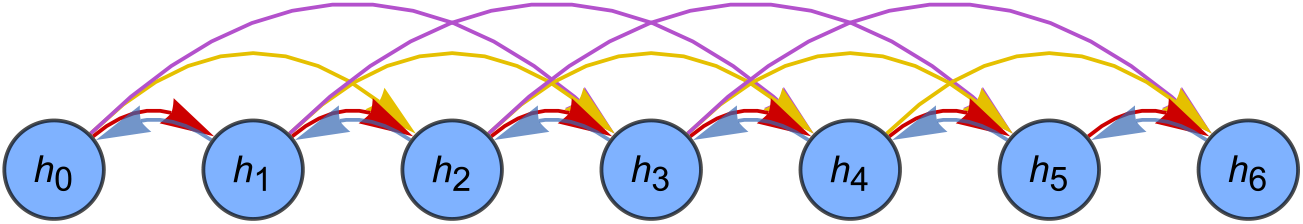
Risk-heterogeneous transmission in SWB. Exposure heterogeneity generates multiple worm-acquisition pathways. Increasing risk produces progressively faster movement through the burden lattice and contributes to the emergence of highly infected hosts.

The corresponding transition operator replaces tridiagonal matrix (2) with Toeplit-type operator

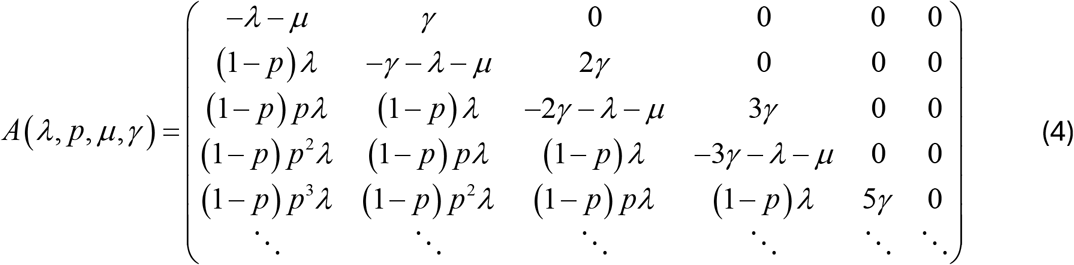

### 3.2 Benchmark Poisson and negative-binomial equilibria

Insight into the model can be obtained in the idealized limit of infinitely many burden strata and no demographic source or turnover,

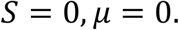

Under homogeneous exposure (*p* = 0), the equilibrium solution of SWB is the Poisson distribution

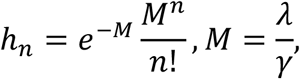

where *M* is the equilibrium mean worm burden.

For heterogeneous exposure (*p* > 0), equilibrium distributions become a risk-weighted mixture of Poisson populations. It assumes a negative-binomial (NB) form with mean burden and aggregation parameters

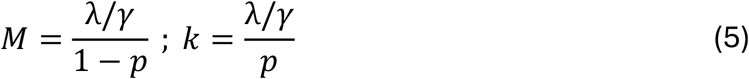

Thus, Poisson and NB distributions emerge naturally as equilibrium states of a common SWB framework. The homogeneous SWB (original version [7-8]) represents the limiting case *p*→0, for which *k*→∞, and the NB distribution converges to Poisson (see Supplement A for details).

Figure 3 shows the resulting family of equilibrium distributions. Increasing risk heterogeneity generates progressively stronger aggregation and broader upper tails while preserving the same underlying transmission structure.

**Figure 3.**
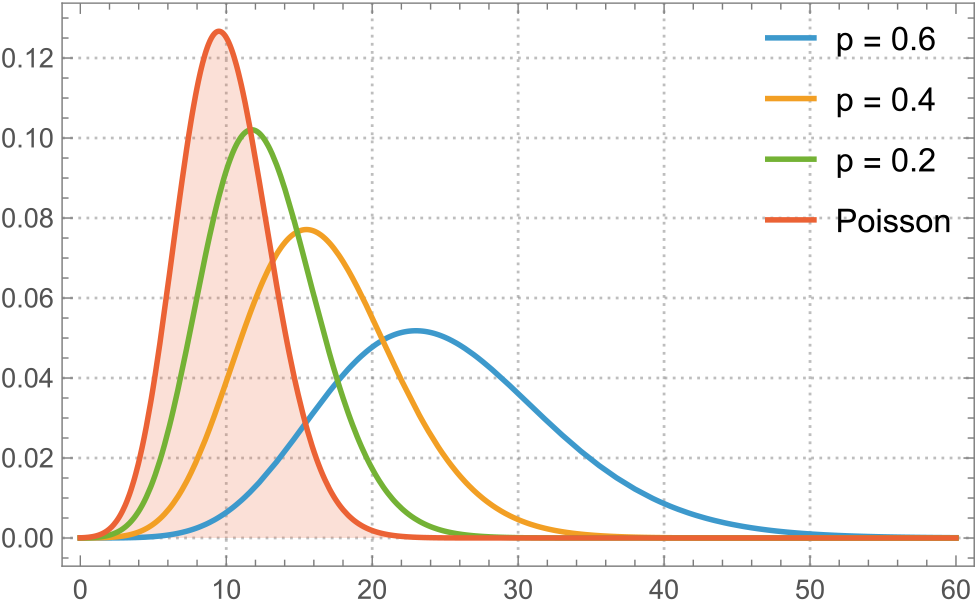
Benchmark equilibrium distributions of the SWB model. Risk heterogeneity generates a family of negative-binomial equilibria NB(λ/p,1-p). As heterogeneity decreases (p→0), the negative-binomial distribution approaches the homogeneous Poisson equilibrium P(λ).

### 3.3 Relation to classical MacDonald (MWB) models

The benchmark equilibria above establish a direct link between SWB and conventional mean worm burden (MWB) theory.

The homogeneous SWB model reproduces the Poisson equilibrium commonly assumed in early MWB formulations, whereas the heterogeneous model generates NB equilibria widely used to represent parasite aggregation. Furthermore, the classical MWB equation is obtained directly as the first statistical moment of the SWB system, as shown previously.

In this sense, Poisson and NB distributions are not independent assumptions but equilibrium consequences of SWB transmission structures.

However, these analytical results rely critically on the absence of demographic turnover. Once infection-free hosts continuously enter the population, the corresponding equilibria generally depart from both benchmark Poisson and NB forms. This distinction and its implications are elaborated below.

For consistency with the transmission models developed below, the mean worm burden *M* will subsequently be denoted by *w*(*t*).

## 4. Coupled Human-Snail Transmission Models for SWB and MacDonald MWB

### 4.1 Human-snail transmission coupling

The stratified worm burden (SWB) models developed in Sections 2-3 are coupled to a snail intermediate-host population through force-of-infection variables acting on humans and snails. Let *z*(*t*) denote the prevalence of infected snails. The human force of infection is *λ*(*t*) = *a z*(*t*), where *a* is the snail-to-human transmission coefficient.

Transmission from humans to snails is mediated by parasite egg release and miracidial stage. Let *E*(*t*) denote the total egg output contributed by the human population. The snail force of infection is assumed proportional to egg release, *Λ*(*t*) = *b E*(*t*), where *b* is the human-to-snail transmission coefficient.

The infected-snail population evolves according to

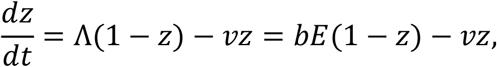

where *v* is snail mortality (2-6/year).

The transmission cycle is represented schematically by *E*→*Λ*→*z*→*λ*. The specific form of *E* depends on the underlying human infection model.

### 4.2 Egg production and mating count

Egg production is assumed proportional to the abundance of mated worm pairs. Let *MC*
denote the expected number of mated worm couples in the human host population. Community egg output is then *E* = *ρ MC*, where *ρ* is the average fecundity of a worm couple. Consequently,

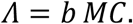

The coefficient *b* combines human-to-snail transmission efficiency and average egg production per worm couple. Unlike previous breakpoint models based on imposed Allee-effect [8], the present framework employs solely the egg-production term *MC*.

For SWB the population mating count is made of mated couples in each stratum *ϕ*_*n*_

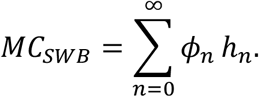

The corresponding MacDonald-type formulation is described by mean worm burden *w*(*t*) and aggregation parameter *k* of underlying negative-binomial distribution *NB*(*w, k*). The expected mating count can be written as

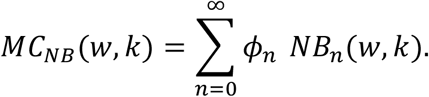

This series can be evaluated in closed form

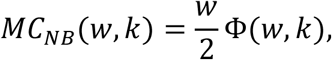

with mating factor *Φ*(*w, k*) (see [3,8], Supplement B).

### 4.3 Coupled human-snail model

For the SWB formulation, dynamic variables *h*_*n*_(*t*) solve a coupled differential system (1) with matrix

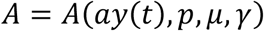

of (4), where human FOI *λ* = *ay*(*t*), depends on infected snail prevalence *y*(*t*) and transmission coefficient *a*.

Substitution *Λ*(*t*) = *b MC*_*SWB*_ yield the snail component of the coupled system

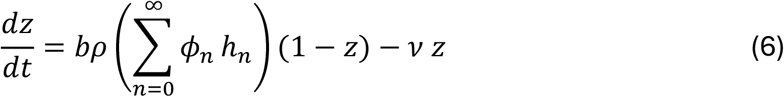

The resulting snail FOI Λ_*NB*_ = *bρ MC*_*NB*_ gives a coupled MacDonald transmission model

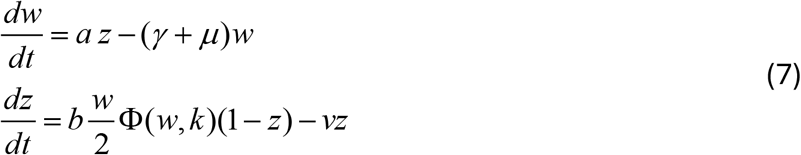

### 4.4 Mass drug administration

MDA is represented as instantaneous event applied at prescribed treatment times *t* = *t*_*j*_.

Let *f* denote treatment coverage and *εε* the fraction of worms surviving treatment (1− ε - clearing efficacy of drug PZQ).

#### SWB implementation

For hosts initially carrying *n* worms, post-treatment worm survival is assumed binomial. The resulting post-treatment burden distribution becomes

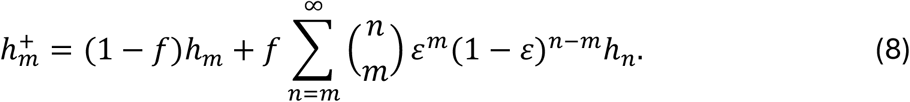

This operator redistributes hosts among burden strata while preserving total population size. This form of MDA is different from earlier versions [7-8,19-23]) and is consistent with NB-assumption of classical MacDonald-type systems.

#### MacDonald implementation

For the MacDonald-NB model, treatment is represented by an instantaneous reduction in mean worm burden,

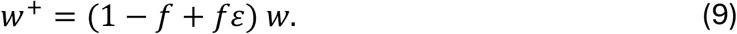

The aggregation parameter *k* is assumed unchanged. Indeed, binomial survival formula (8) applied to NB(w,k) would change its mean according to (9), and preserve k.

Both formulations therefore achieve the same expected proportional reduction in mean worm burden, while differing in their treatment-induced changes of the underlying burden distribution. It has significant implications for MDA of SWB system vs. NB-type MacDonald.

## 5. Demographic SWB equilibria

### 5.1 Recruitment of infection-free hosts

Real host populations are continually renewed through birth, maturation, and mortality. These demographic processes introduce source terms into the SWB equations, yielding equilibrium conditions

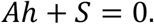

For child populations, the demographic source consists primarily of newborn hosts entering the infection-free stratum,

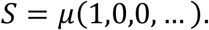

Such recruitment cannot be represented naturally within classical MWB formulations because MWB tracks only the average worm burden. In contrast, SWB explicitly follows the entire burden distribution and accommodates demographic inflow through specified burden strata.

This seemingly simple distinction has important consequences for endemic equilibria, transmission dynamics, and model calibration.

### 5.2 Child populations

Figure 4 shows equilibrium burden distributions for several demographic turnover rates. When turnover is weak, demographic recruitment has little effect and SWB equilibria remain close to their corresponding Poisson or NB benchmark distributions. However, as turnover approaches realistic values for childhood populations, marked departures emerge. The largest deviations occur in the lowest burden strata. Continuous introduction of infection-free newborns produces excess density at low *n* = 0,1,2,…while simultaneously reducing the relative weight of intermediate infection classes. As a result, the equilibrium distribution ceases to follow either a Poisson or NB shape despite identical transmission parameters.

**Figure 4.**
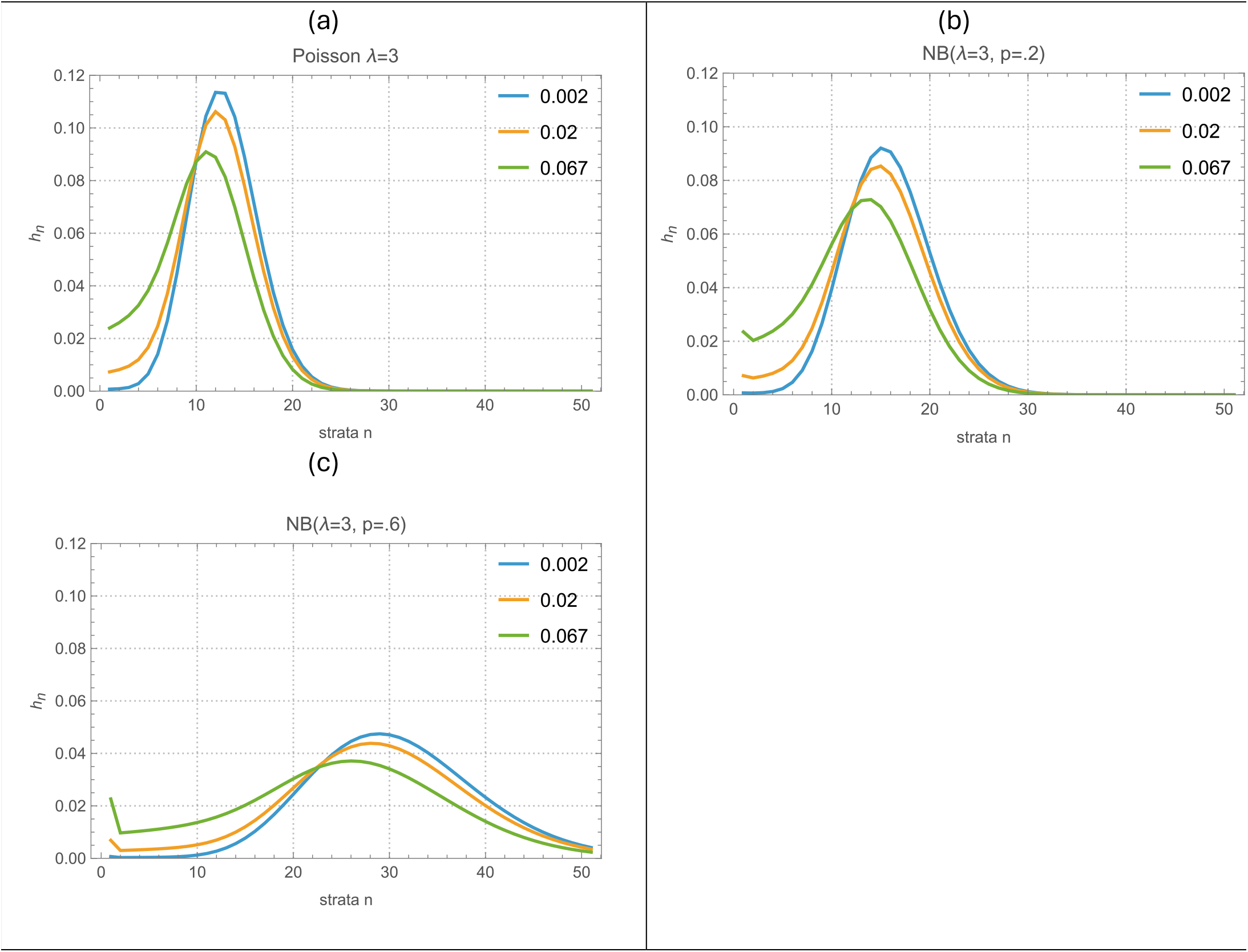
Effects of demographic turnover on equilibrium burden distributions. Increasing demographic turnover (µ) shifts equilibrium densities toward lower burden classes through continual recruitment of infection-free hosts. As µ→0, solutions approach the benchmark Poisson or negative-binomial equilibria.

These effects become increasingly pronounced as demographic turnover increases.

#### 5.3 Coupled child-adult populations

The influence of demographic recruitment extends beyond children. Adult populations inherit infection through recruitment from maturing child cohorts rather than directly from newborn hosts.

Figure 5 compares equilibrium distributions in a coupled child-adult system. Child equilibria depart substantially from their corresponding NB benchmark because demographic input is concentrated in infection-free hosts. Adult equilibria remain closer to the classical NB prediction because their source population is already partially infected.

**Figure 5.**
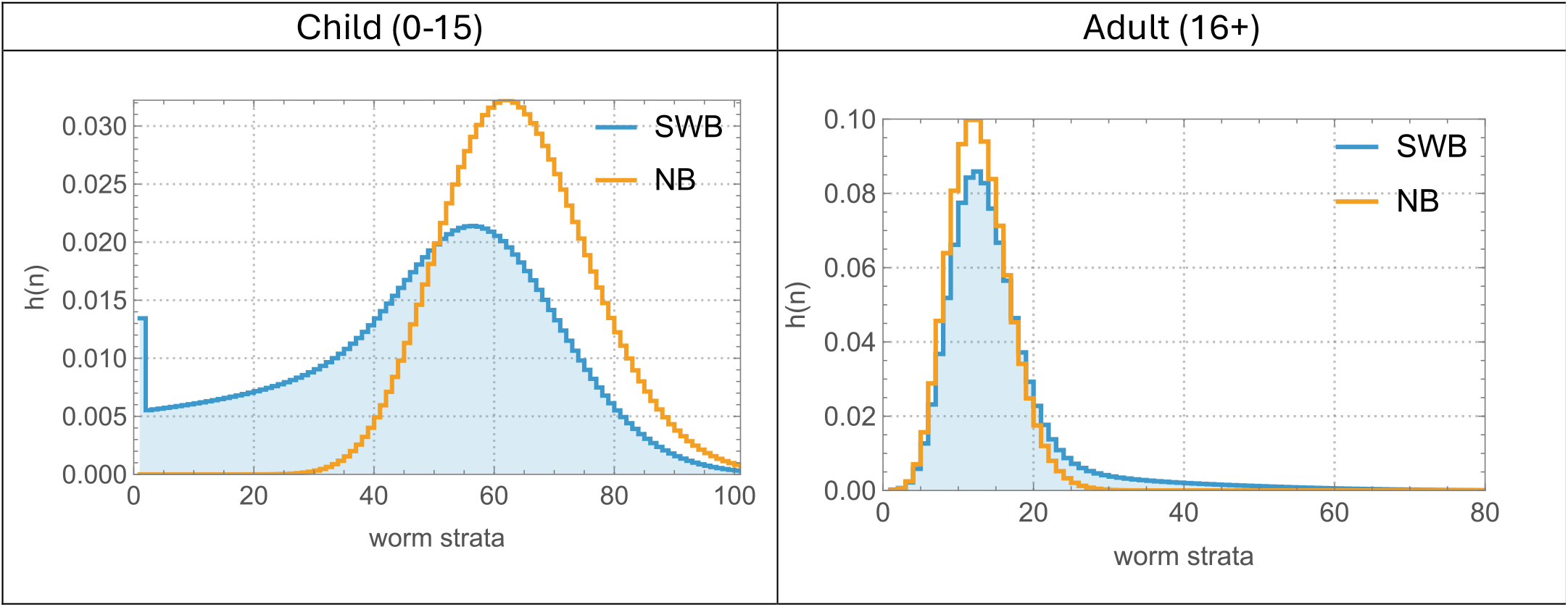
Equilibrium distributions in coupled child-adult populations. Child distributions exhibit substantial departures from benchmark NB equilibria owing to recruitment of infection-free newborns. Adult distributions remain closer to benchmark expectations because recruitment occurs through partially infected maturing hosts.

This difference illustrates how demographic structure shapes endemic burden distributions even when transmission parameters remain unchanged.

### 5.4 Departure from Classical Poisson/NB equilibria

The preceding examples demonstrate that demographic recruitment fundamentally alters the structure of endemic equilibria. Consequently, classical Poisson and NB distributions in MacDonald-type models should be viewed as analytical benchmarks rather than realistic endemic states.

The discrepancy becomes increasingly important at moderate and high transmission intensities. Figure 6 compares equilibrium predictions obtained from demographic SWB and classical NB formulations over a range of transmission and aggregation parameters.

**Figure 6.**
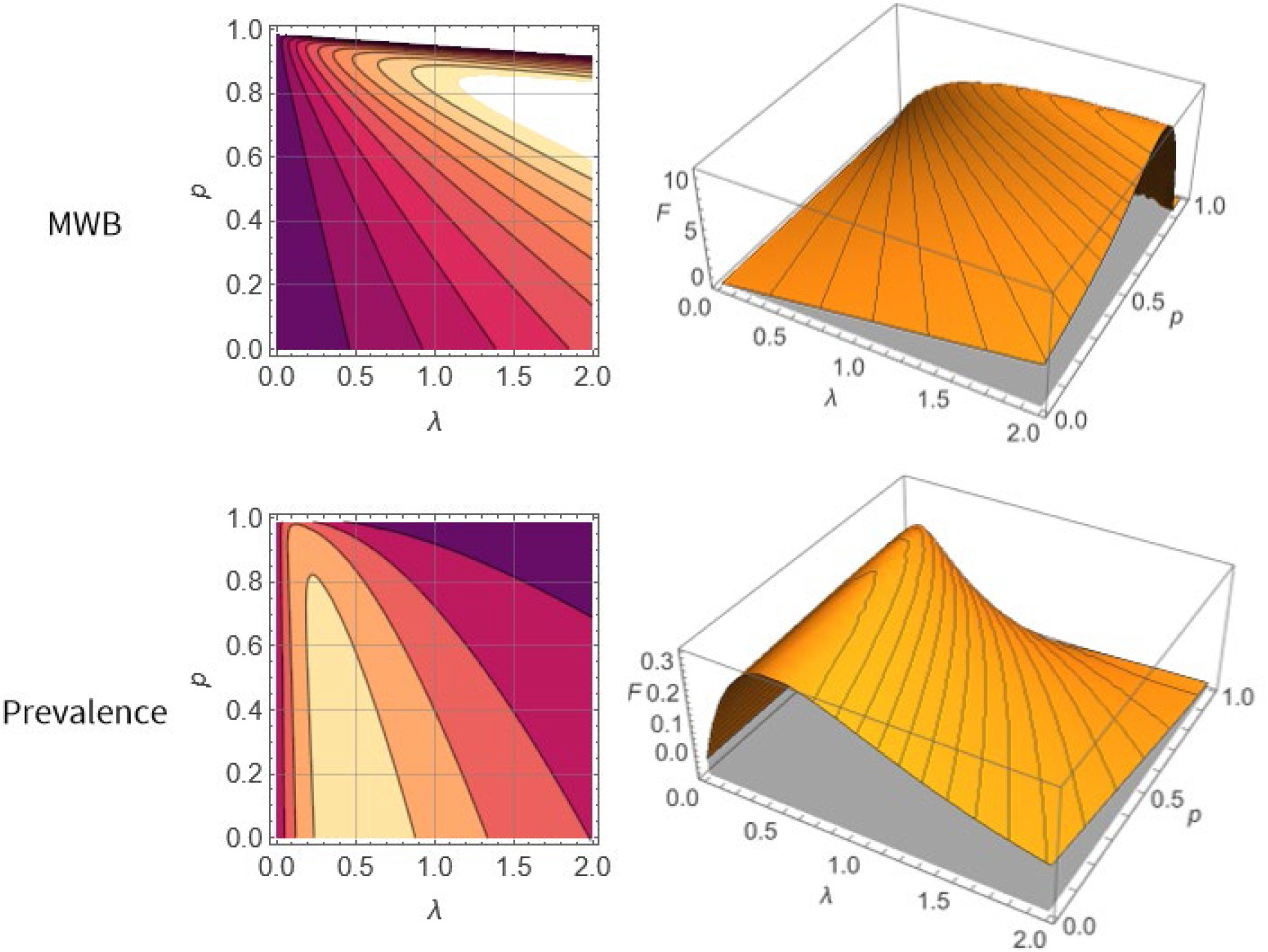
Demographic SWB versus classical Poisson/NB equilibria. Surface plots show differences between demographic SWB predictions and corresponding benchmark Poisson or negative-binomial equilibria across transmission and aggregation SWB parameters (λ, p), for MacDonald system we use benchmark equilibrium values (M, k) derived from (λ, p) (equation (5) Top panels do it for MWB the lower panels repeat it for prevalence. Right and left panels offer two different views of the difference functions (contours vs. 3D surface plot). Discrepancies increase with transmission intensity and affect mean worm burden, worm prevalence, and mated-couple prevalence.

Differences appear not only in the burden distribution itself but also in derived quantities such as mean worm burden, worm prevalence, and mated-couple prevalence. Notably, substantial discrepancies occur within the transmission range occupied by many observed SCORE communities.

### 5.5 Implications for transmission modeling

The effects of demographic sources extend beyond equilibrium calculations.

First, calibration based solely on Poisson or NB assumptions may lead to biased estimates of transmission intensity because the observed burden distribution includes demographic contributions that are absent from classical MWB theory.

Second, demographic source terms influence dynamic responses to treatment and transmission reduction by continuously replenishing low-burden host classes.

Finally, age-structured populations cannot generally be represented by a single equilibrium Poisson or NB distribution, since different demographic processes operate across age groups.

For these reasons, demographic SWB provides a more realistic framework for endemic parasite populations than classical MWB models while retaining Poisson and NB distributions as useful limiting cases and analytical benchmarks.

## 6. Coupled Human-Snail Systems

To investigate endemic transmission and elimination thresholds, we couple the human infection models developed above with a dynamic snail population. Let *z*(*t*)denote infected-snail prevalence and assume human force of infection is proportional to snail infection, *λ* = *az*, where *a* is the snail-to-human transmission coefficient.

For the SWB formulation, human contamination of the environment (snail force of infection) is determined by the density of mated worm pairs,

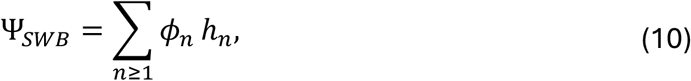

where *ϕ*_*n*_ counts the expected number of mating pairs in stratum *h*_*n*_ (Supplement B). Hence, snail FOI

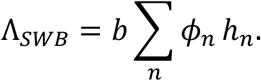

For the MWB (MacDonald) formulation, the density of mated pairs can be estimated as in (10), where strata variables {*h*_*n*_} are given by *NB* (*w, k*). The net result is

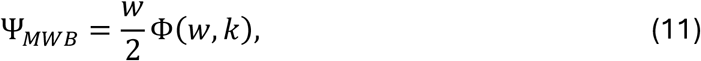

with mating factor Φ(*w, k*) expressed though a hypergeometric function (Supplement B). Ther corresponding FOI takes the form

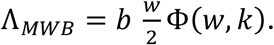

In both formulations snail infection evolves according to SI-model with susceptible fraction (1-z), infected z, the snail FOI Λ,

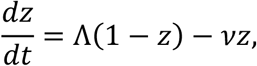

where *v* is snail mortality.

The two models share an identical snail component and differ only in the representation of the human contribution Λ to transmission.

### 6.1 Equilibria and Stability Analysis

A human endemic state for *λ* = *az*, can be expressed through variable z. For MacDonald

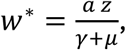

for SWB we get its equilibrium distribution *h*^∗^(*az, p, γ*, µ), expressed through z. Substitution into snail FOI Λ(w^∗^) or Λ (*h*^*^) gives a reduced one-dimensional z-equation

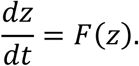

For the SWB model,

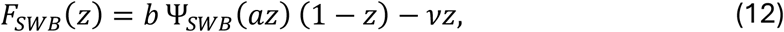

where Ψ_*SWB*_(*az*) is obtained from the equilibrium SWB distribution *h*^∗^(*az, p, γ*, µ) via equation (10). Similarly,

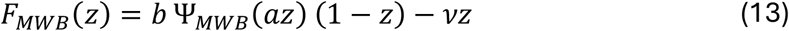

with factor Ψ_*MWB*_ given by (11).

Equilibria of the reduced and fully coupled model correspond to zeros of *F*(*z*) = 0, Their stability is determined by *F*^′^(*z*^∗^). A stable equilibrium satisfies *F*^′^(*z*^∗^) < 0, whereas unstable equilibria satisfy *F*^′^(*z*^∗^) > 0.

The disease-free state *z* = 0 always exists. Depending on model parameters (*a, b)*et al, additional positive equilibria may arise.

The analysis involved numerical exploration of function *F* (*z, a, b*,…), its equilibria *z*^∗^(*a, b*, …); local maxima (for breakpoint analysis); and for SWB ‘zero stability’-Jacobian matrix analysis.

Three qualitatively distinct regions were identified:

- Disease-free (zero) region: only *z* = 0 exists and is stable.
- Breakpoint (bistable) region: stable zero and endemic equilibria coexist, separated by an unstable breakpoint.
- Single-endemic region: only one positive stable equilibrium exists.

To illustrate these cases, Figure 7 shows representative plots of *F*(*z*) for both SWB and MWB systems. Intersections with the horizontal axis correspond to equilibria, while the direction of the vector field is determined by the sign of *F*(*z*).In the disease-free regime, *F*(*z*) crosses zero only at *z* = 0. In the breakpoint regime, *F*(*z*)crosses the axis three times, producing the classical bistable structure. In the single-endemic regime, the disease-free equilibrium is unstable and all trajectories converge to a positive endemic equilibrium.

### 6.2 Stability regions in (*a, b*)-space

To determine where each transmission regime occurs, we numerically evaluated the reduced equation

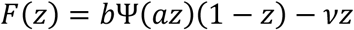

over a range of transmission parameters (*a, b*).

**Figure 7.**
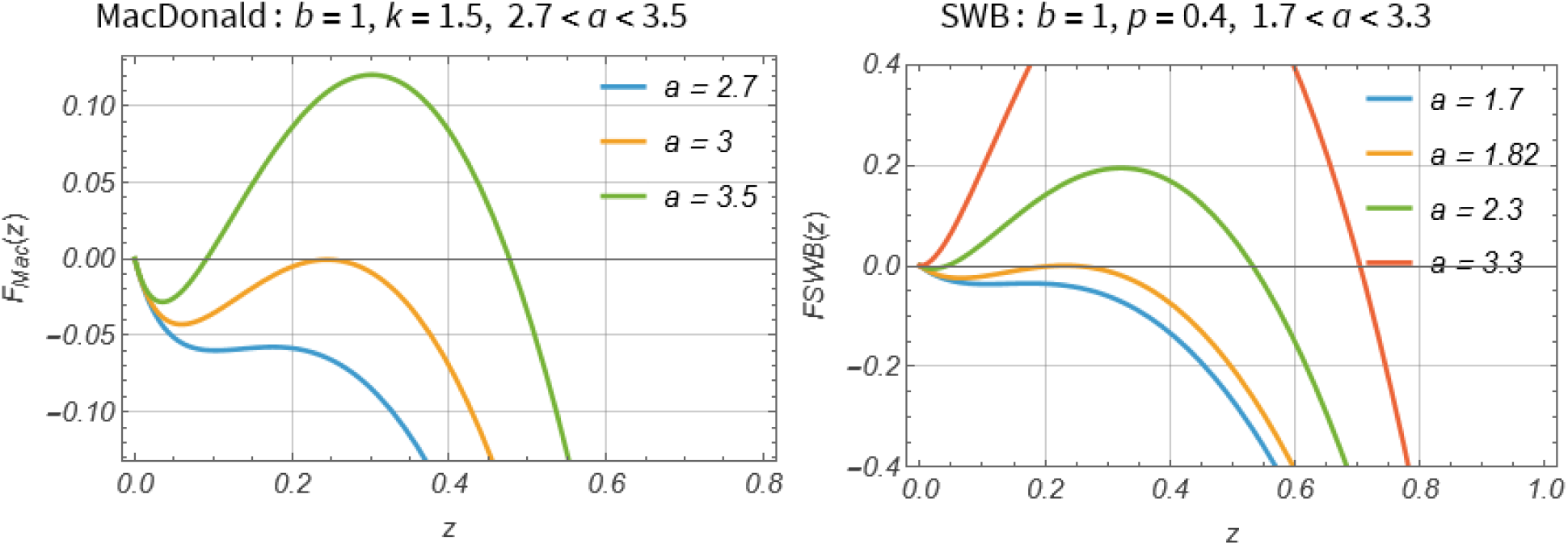
Representative plots of the reduced snail equation F(z) = bΨ(az)(1 − z) − vvz. Zeros of F(z)correspond to equilibria of the coupled system. Panels illustrate the three qualitative regimes identified in the stability analysis: disease-free transmission, bistability with an unstable breakpoint, and a single endemic equilibrium. Solid circles denote stable equilibria and open circles denote unstable equilibria. Corresponding SWB and MWB examples are shown for parameter values selected from the indicated regions of the (a, b)-plane.

We fixed parameter values 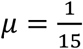, 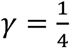 throughout this section, also dispersal *p* = 0.4 for SWB, and aggregation k=1.5 for MacDonald.

The resulting stability regions are shown in Figure 8. For SWB, we derived two bifurcation curves partitioning the (*a, b*)-plane into three regions:

i. Stable disease-free (zero) equilibrium;
ii. bistability with a transmission breakpoint;
iii. stable endemic equilibrium

**Figure 8.**
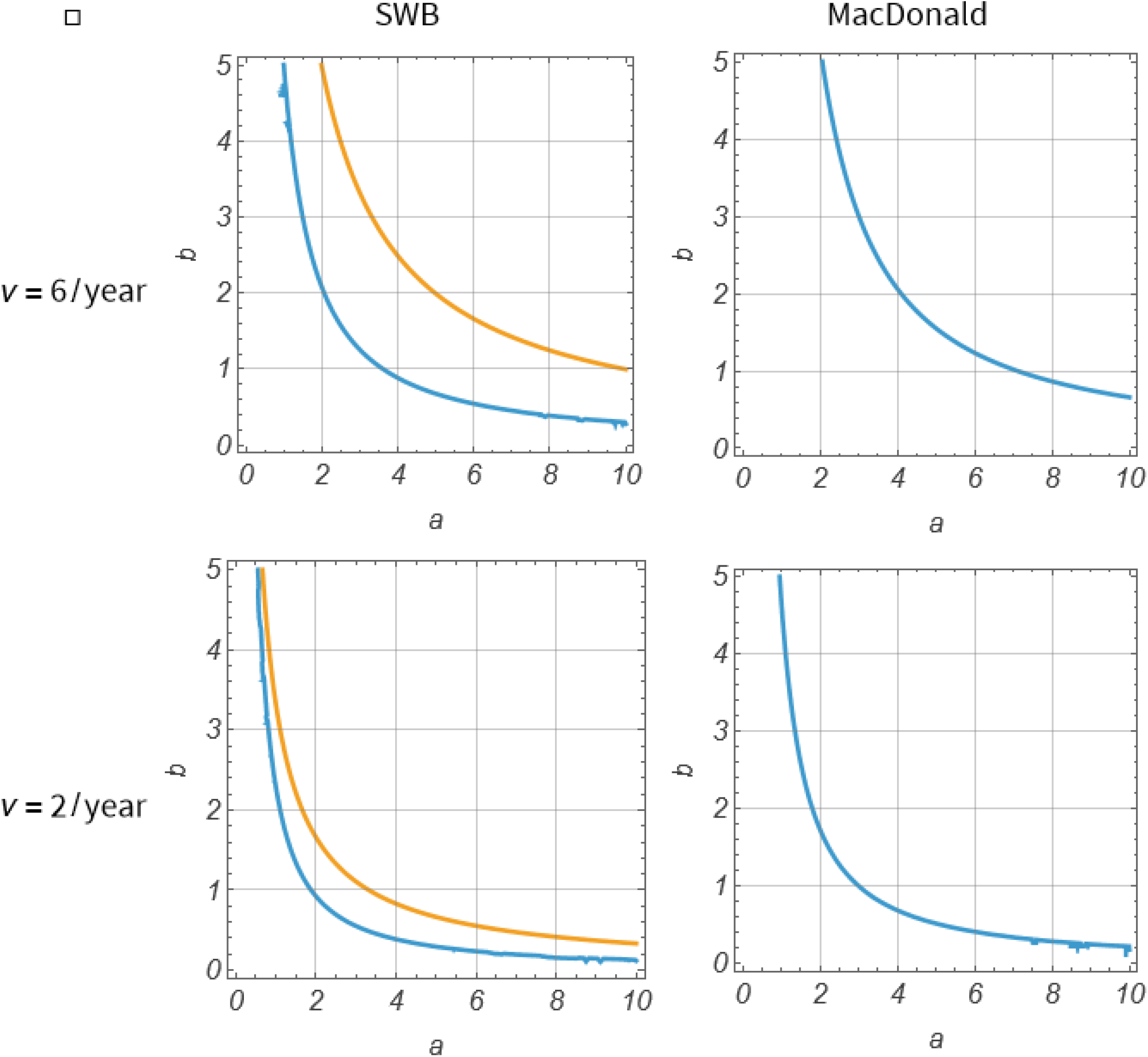
Stability regions of the coupled human-snail system in the (a, b) parameter plane. Regions correspond to (i) stable zero (disease-free) equilibrium (below blue curves) for both SWB and MacDonald; (ii) bistability (breakpoint regime), for SWB it consist of the band between blue and yellow curves, for MacDonald – all points above blue curve; (iii) stable endemic and unstable zero: all points above yellow curve. SWB model (p = 0.4). Right: MacDonald model (k=1.5). The bistable region is substantially larger in the MWB formulation, indicating a broader range of transmission conditions under which elimination remains theoretically attainable.

The upper boundary (yellow curve) corresponds to loss of stability of the disease-free (zero) equilibrium. The lower boundary (blue curve) corresponds to a pitchfork bifurcation ‘stable zero’ to ‘bistable triplet’, same as MacDonald blue curve.

Function *F*(*z*) and its (*a,b*)-stability regions depend on other model inputs, including snail mortality *v*. Here we demonstrated 2 choices: short-lived snails *v* = 6 (upper panels of Figure 8) vs long lived opnes *v* = 2 (lower panels). Reduced snail mortality pushes bifurcation curves lower.

The SWB and MWB diagrams exhibit similar qualitative structure but differ quantitatively. In particular, the bistable region is substantially smaller in the SWB model. As a result, parameter combinations that admit a transmission breakpoint in the MWB model could correspond to a single endemic equilibrium in the SWB formulation.

This distinction reflects the different descriptions of mating structure. In the MWB model, transmission is represented through the negative-binomial mating function Φ(*w, k*). In the SWB formulation, transmission is computed directly from the equilibrium burden distribution and therefore retains information on worm aggregation across host classes.

Consequently, the calibrated MWB model generally predicts a broader parameter domain in which transmission interruption is achievable.

The bifurcation structure shown in Figure 8 provides the basis for the intervention experiments of Section 7.

## 7. MDA Analysis and Comparison

We compare MDA outcomes predicted by the SWB and NB-MacDonald formulations under both externally specified and transmission-coupled force of infection scenarios.

Throughout this section we consider annual treatment of the child population (0-15 years), demographic turnover 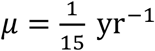 and worm mortality 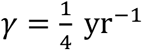

Treatment is characterized by coverage *f* and surviving worm fraction *ε*. Unless otherwise stated,

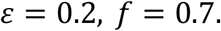

based on Praziquantel clearing efficacy (80%), and typical coverage.
For the SWB model, MDA acts through an instantaneous redistribution among burden strata according to binomial worm survival,

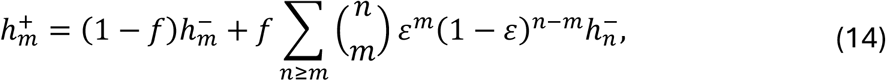

whereas for the MacDonald model

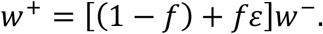

The aggregation parameter *k* stays fixed before and after MDA, consistent with reshuffle

(14) applied to the underlying NB distribution.

To facilitate comparison we record two summary outputs: *w*(*t*) −mean worm burden, and *P*_*c*_(*t*) -observable prevalence based on hosts carrying at least one mated worm pair. For

SWB,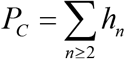, while NB-MacDonald has *PC* expressed through its mean and aggregation,

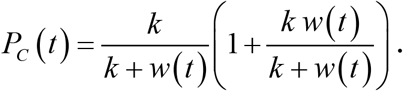

### 7.1 Fixed Force of Infection

We first consider a hypothetical setting where human exposure remains constant and is unaffected by local transmission. Such a scenario may arise when several communities share common water-contact sites so that local reductions of infection in one of them has negligible effect on the overall cercarial environment.

The SWB parameters are fixed at *λ* = 1.5, *p* = 0.4. Two MDA comparisons are considered.

Case 1: Identical forces of infections (*λ* = 1.5) are imposed on both models, along with µ, *γ*. For the MacDonald formulation, equilibrium matching gives

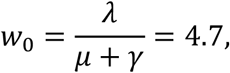

To pair the benchmark MacDonald NB to SWB (*λ, p*) with *p* = 0.4 we take

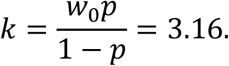

Case 2: Calibrated MacDonald Model

The MacDonald parameters are calibrated to reproduce the endemic SWB equilibrium for

(*λ, p*) = (1.5,0.4),

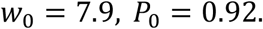

This yields

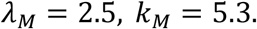

significantly higher than SWB values (*λ*_*M*_ > *λ*_*SWB*_).

In both cases repeated treatment rapidly approaches a periodic limit cycle. Since transmission is maintained externally, neither system admits elimination. After each treatment round worm burden decreases sharply, followed by recovery toward the periodic attractor. Consequently, MDA alone cannot interrupt transmission under fixed FOI; additional interventions such as WASH or environmental control would be required to reduce the underlying exposure rate.

**Figure 9.**
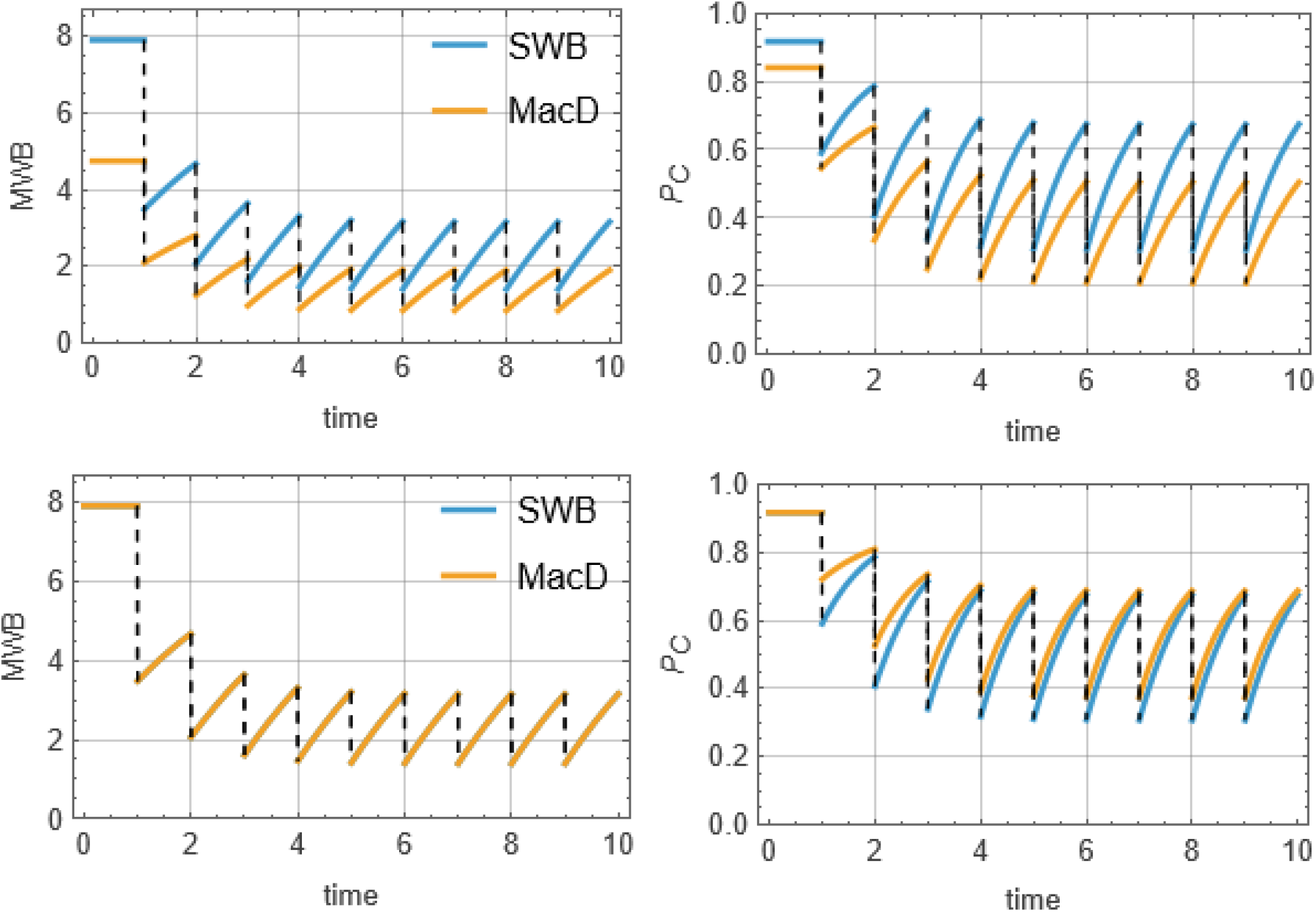
MDA histories under fixed force of infection. Top row: identical environmental input (λ = 1.5) imposed on both SWB and MacDonald models. Bottom row: calibrated MacDonald model reproducing the endemic SWB equilibrium (w_0_ = 7.9, P_0_ = 0.92).Left panels show mean worm burden w(t); right panels show prevalence P_c_(t). In the identical-FOI comparison, MacDonald equilibrium levels and rebound are substantially lower than those of SWB. Calibration greatly improves agreement, although rebound remains somewhat stronger for SWB. Both systems converge to stable periodic MDA cycles and neither can achieve elimination under fixed FOI.

### 7.2 MDA in Coupled Human-Snail Systems

We next consider the full transmission cycle in which human and snail infection evolve jointly according to human FOI *λ* = *az*, and snail equation

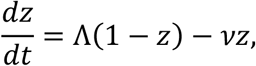

With snail FOI given by

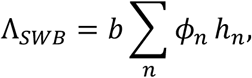

and

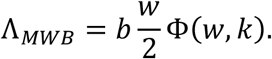

The transmission parameters used for MDA experiments were selected from the stability diagrams of Section 6. Specifically, we chose parameter values lying within the single endemic equilibrium region of the SWB model, above the endemic threshold and outside the SWB bistable regime. This choice ensures that elimination cannot occur through purely local stability effects and therefore provides a stringent comparison of rebound behavior.

As above two cases are examined.

Case 1: Identical Environmental Inputs. Both models use (*a, b*) = (4, 1), with SWB *p* = 0.4 and MacDonald *k* = 0.5.

Case 2: Calibrated MacDonald Model

The MacDonald parameters are adjusted to reproduce the SWB endemic equilibrium. Matching endemic MWB and prevalence yields

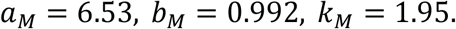

which leaves b intact but raises markedly (*a,k)*.

Under repeated MDA, all state variables *w*(*t*), *P*_*c*_(*t*), *z*(*t*) decline simultaneously:

Initially the trajectories appear similar, particularly in the calibrated case. However, differences become increasingly apparent near the transmission threshold. Rebound of worm burden, prevalence, and snail infection is consistently stronger in the SWB system. Continued treatment drives the calibrated MacDonald trajectory toward its breakpoint, whereas the corresponding SWB trajectory remains within the basin of attraction of the endemic equilibrium and ultimately approaches a nonzero periodic orbit.

**Figure 10.**
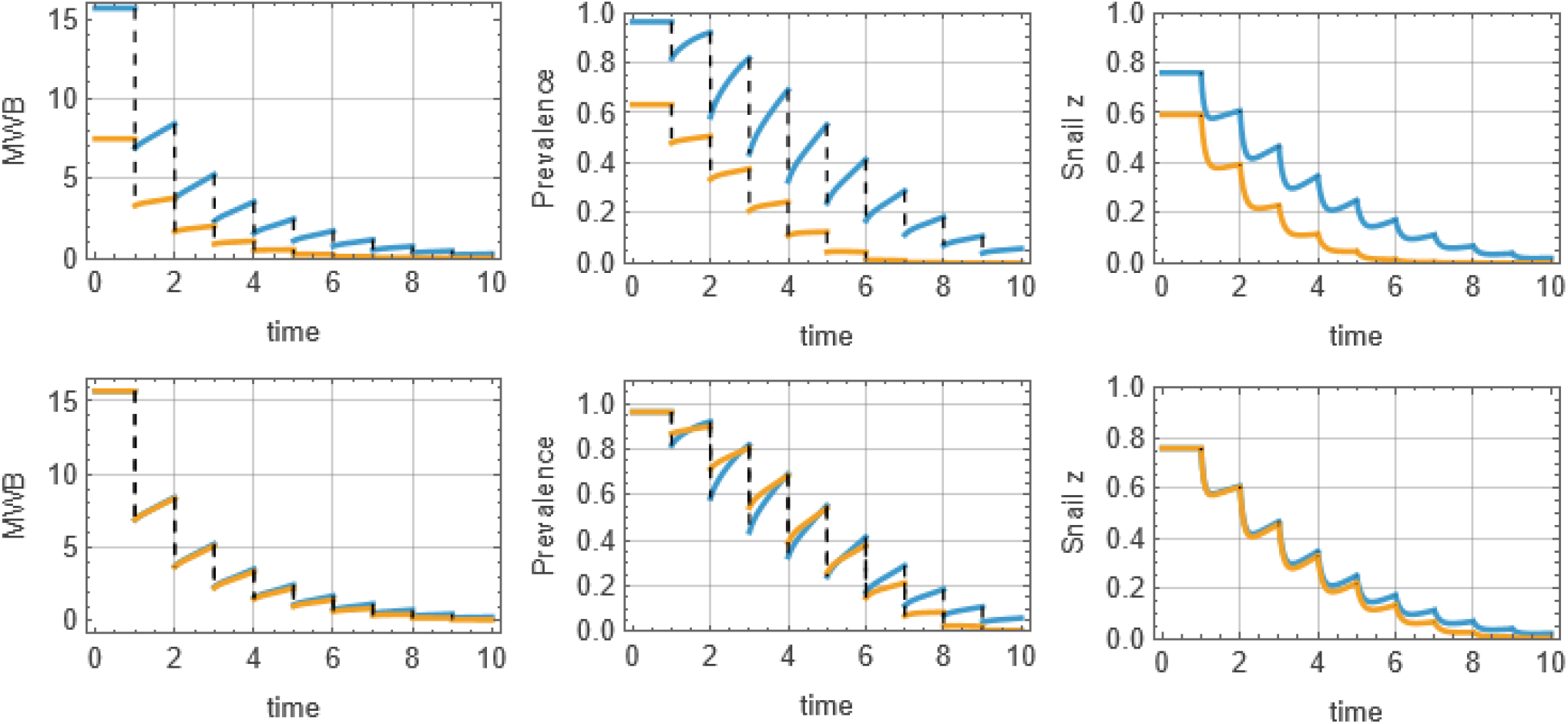
MDA histories for coupled human-snail systems. Top row: identical transmission environment (a, b) = (4, 1) for SWB (blue line) and MacDonald (yellow). Bottom row: calibrated MacDonald model (a, b, k) = (6.53, 0.992, 1.95) reproducing the endemic SWB equilibrium. Columns show mean worm burden w(t), prevalence P_c_(t), and infected-snail prevalence z(t). Identical environmental inputs produce substantially smaller endemic levels and weaker rebound in the MacDonald model. Calibration improves agreement; nevertheless SWB consistently exhibits stronger rebound. Extension of the simulations shows that the calibrated MacDonald system eventually crosses its breakpoint and collapses toward elimination, while the corresponding SWB system approaches a persistent limit cycle

### 7.3 WHO Elimination Target and Post-Treatment Rebound

Current WHO guidelines focus on prevalence-based targets rather than direct assessment of transmission breakpoints. To evaluate the implications of this strategy, we consider a simple stopping rule:

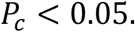

MDA is terminated once prevalence falls below 5.

The initial treatment trajectories are very similar in both models. In the calibrated simulations the prevalence threshold is reached after approximately nine annual treatment rounds. At this stage the MWB, prevalence, and snail prevalence appear comparable, suggesting successful control.

However, subsequent dynamics diverge dramatically.

In the MacDonald model the system has crossed its transmission breakpoint. Following cessation of treatment all state variables continue to decline and converge to the disease-free equilibrium,

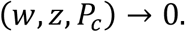

In contrast, the SWB system has no breakpoint. After treatment stops, both human and snail infection recover and eventually return to the endemic attractor.

Thus identical prevalence targets can correspond to fundamentally different dynamical states.

These simulations highlight a central conclusion of this study: models calibrated to reproduce the same endemic prevalence and mean worm burden can nevertheless generate sharply different predictions regarding breakpoints, elimination thresholds, and post-MDA rebound.

**Figure 11.**
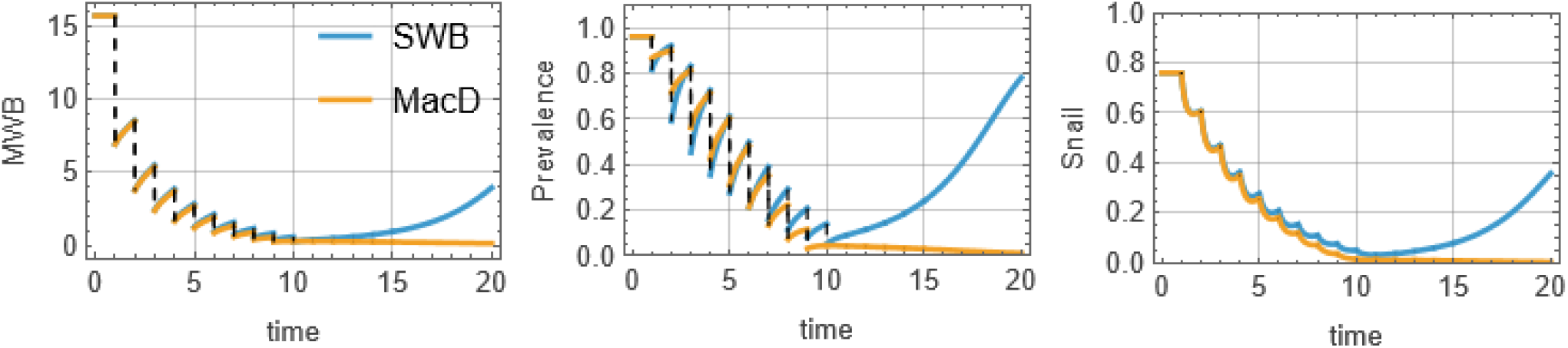
Divergent post-treatment dynamics after reaching the WHO prevalence target P_c_ < 0.05. MDA is discontinued once prevalence falls below 5%. Panels show mean worm burden w(t), prevalence P_c_(t), and infected-snail prevalence z(t). Initial treatment histories are nearly indistinguishable. Following cessation of treatment, however, the calibrated MacDonald system lies below its transmission breakpoint and converges to the disease-free equilibrium, whereas the SWB system remains above its breakpoint and rebounds toward endemic transmission. This illustrates that prevalence-based control targets do not necessarily imply elimination of transmission.

## 8. Conclusions

The stratified worm-burden framework developed here provides a direct bridge between classical macroparasite theory and burden-structured population dynamics. We show that the Poisson and negative-binomial distributions widely used in MWB models emerge naturally as equilibrium solutions of idealized SWB systems, giving these familiar statistical descriptions a mechanistic foundation.

Incorporation of demographic recruitment and host turnover fundamentally modifies this picture. The resulting endemic burden distributions differ systematically from their Poisson and NB benchmarks, demonstrating that classical distributions should be viewed primarily as limiting reference states rather than realistic demographic equilibria. This distinction becomes important when models are calibrated to endemic prevalence and mean worm burden but differ in their underlying burden structure.

These findings indicate that demographic recruitment is not a minor correction but a structural component of endemic infection dynamics.

For coupled human-snail transmission systems, we develop a consistent mating-based transmission formulation in which egg production is determined directly from the worm-burden distribution. In contrast to previous SWB breakpoint studies, breakpoint behavior is obtained without imposing additional Allee-type transmission assumptions. The analysis reveals that transmission breakpoints are not universal properties of burden-structured systems. Instead, bistability occurs only within a restricted region of transmission parameter space, whereas corresponding NB-MacDonald models exhibit breakpoint behavior over a much broader range of conditions.

A particularly important finding concerns transmission breakpoints. In contrast to classical negative-binomial MacDonald formulations, where breakpoint behavior occurs over broad parameter ranges, the SWB framework restricts bistability to a relatively narrow domain of transmission conditions. Outside this domain the system possesses a single endemic attractor. Thus breakpoint behavior is not a generic consequence of worm aggregation or mating effects, but rather a contingent dynamical property whose existence depends on the underlying burden structure and demographic processes.

These differences have direct implications for evaluating elimination programs. Even when calibrated to the same endemic prevalence and mean worm burden, SWB and MacDonald models can produce markedly different predictions for post-MDA rebound, persistence, and long-term elimination. Reliance on prevalence targets alone may therefore provide an incomplete assessment of interruption of transmission. So morbidity control does not necessarily imply interruption of transmission, and prevalence alone may provide a misleading indicator of elimination success.

More broadly, the SWB framework demonstrates that explicit representation of worm-burden distributions can unify endemic equilibria, demographic turnover, transmission dynamics, and intervention analysis within a single mathematical structure. By linking classical macroparasite theory to burden-resolved population dynamics, the framework offers an alternative foundation for evaluating parasite control and elimination strategies.

## Supporting information

Technical Supplements

## Data Availability

All data and computer codes produced in the present study are available upon reasonable request to the authors

