## Supplementary material for "A Stratified Worm Burden Framework for Endemic Equilibria, Demography, and Mass Drug Administration: Linking Classical Macroparasite Models to Transmission Dynamics": Technical Supplements

### Supplement A. Benchmark Poisson and Negative-Binomial Equilibria of the SWB Model

#### S1.1 Homogeneous SWB equilibrium

Consider the idealized SWB system with infinitely many burden strata, no demographic recruitment, and no host turnover ($\mu = 0$),

$$\frac{dh}{dt} = Ah,$$

where

$$h = (h_0, h_1, h_2, \dots)^T$$

and the transition rates are generated by worm acquisition rate $\lambda$ and per-worm mortality rate $\gamma$.

At equilibrium,

$$Ah = 0.$$

The balance equations are

$$0 = \lambda h_{n-1} + \gamma(n+1)h_{n+1} - (\lambda + \gamma n)h_n, n \geq 0,$$

with $h_{-1} = 0$.

Define the probability generating function

$$G(z) = \sum_{n=0}^{\infty} h_n z^n.$$

Multiplying the balance equation by $z^n$and summing over all $n$yields

$$0 = \lambda(z-1)G(z) + \gamma(1-z)G'(z),$$

or equivalently

$$G'(z) = \frac{\lambda}{\gamma}G(z).$$

Solving,

$$G(z) = C\exp\left(\frac{\lambda}{\gamma}z\right).$$

Using the normalization condition $G(1) = 1$,

$$C = \exp\left(-\frac{\lambda}{\gamma}\right).$$

Thus

$$G(z) = \exp\left[\frac{\lambda}{\gamma}(z-1)\right].$$

This is the generating function of a Poisson distribution with mean

$$M = \frac{\lambda}{\gamma}.$$

Therefore

$$h_n = e^{-M}\frac{M^n}{n!}, n = 0,1, \ldots$$

is a null-vector of the homogeneous SWB operator $A$.

Hence the classical Poisson worm distribution emerges as the equilibrium state of homogeneous SWB.

---

#### S1.2 Risk-heterogeneous SWB equilibrium

Suppose host exposure is distributed among risk classes $r$with weights

$$w_r = (1-p)p^r, r = 0,1,2, \ldots$$

and acquisition rates

$$\lambda_r = \lambda q_r.$$

Within each risk class, equilibrium worm burdens remain Poisson,

$$h_n^{(r)} = e^{-M_r}\frac{M_r^n}{n!},$$

with class-specific mean

$$M_r = \frac{\lambda_r}{\gamma}.$$

The population distribution is therefore a mixture

$$h_n = \sum_r w_r \, h_n^{(r)}.$$

The corresponding generating function is

$$G(z) = \sum_r w_r \exp\ [M_r(z-1)].$$

For the geometric risk spectrum used in the main text, the sum can be evaluated explicitly and gives

$$G(z) = \left[1 + \frac{M}{k}(1-z)\right]^{-k},$$

where $M$is the population mean worm burden and $k$is an aggregation parameter determined by the risk distribution.

This is precisely the generating function of the negative-binomial distribution, of mean M and aggregation k

$$h_n = \frac{\Gamma(n+k)}{\Gamma(k)\Gamma(n+1)} \left(\frac{k}{k+M}\right)^k \left(\frac{M}{k+M}\right)^n.$$

Thus the NB distributions arise as null-vector of the heterogeneous SWB operator.

---

**S1.3 Poisson limit**

The NB generating function,

$$G(z) = \left[1 + \frac{M}{k}(1-z)\right]^{-k},$$

satisfies

$$\lim_{k\to\infty} G(z) = \exp[M(z-1)],$$

recovering the Poisson generating function.

Accordingly,

$$\lim_{k\to\infty} NB(M,k) = Poisson(M).$$

The homogeneous SWB model therefore represents a limiting case of the broader risk-heterogeneous framework.

#### S1.4 Interpretation

The generating-function derivation reveals that Poisson and negative-binomial worm distributions are not imposed statistical assumptions. Rather, they arise as equilibrium null-vectors of idealized SWB transition operators.

The Poisson equilibrium corresponds to homogeneous exposure and worm loss, whereas the negative-binomial equilibrium results from superposition of multiple exposure-risk classes. Both solutions require the absence of demographic source terms. Once recruitment and turnover are introduced, the equilibrium condition becomes

$$Ah + S = 0,$$

and the resulting endemic distributions generally depart from the classical Poisson and negative-binomial families.

#### Supplement B: Mated count for SWB and NB-MacDonald

Mated count for stratum $h_n$ (hosts carrying n worms) can be estimated assuming binomial male-female distribution with sex ratio 1:1 [May, 1977]. It gives

$$\phi_n = \frac{n}{2}\left(1 - 2^{-n}\binom{n}{n/2}\right); n \geq 2,$$

and the population mated count $\sum_{n\geq 2} \phi_n h_n$.

For MacDonald-type system strata variables $\{h_n\}$ are distributed according to $NB(w,k)$, and the combined mated count

$$MC_{Mac}(w,k) = \sum_{n\geq 2} \phi_n h_n(w,k)$$

can be evaluated in closed form through hypergeometric function ${}_2F_1(a,b,c \mid z)$, Specifically,

$$MC_{Mac}\left(w,k\right)=\frac{w}{2}\,{}_2F_1\left(\frac{1}{2},1+k,2\middle|-\frac{2w}{k}\right)$$

([May, 1977], [Gurarie-King 2014].
